# Association of Cut-point Free Metrics and Common Clinical Tests among Older Adults after Proximal Femoral Fracture

**DOI:** 10.1101/2025.02.13.25322210

**Authors:** Hananeh Younesian, David Singleton, Beatrix Vereijken, Judith Garcia-Aymerich, Lynn Rochester, Martin Aursand Berge, Monika Engdal, Joren Buekers, Sarah Koch, Jorunn L. Helbostad, Paula Alvarez, Carl-Philipp Jansen, Kamiar Aminian, Anisoara Ionescu, Clemens Becker, Brian Caulfield, the Mobilise-D Consortiumon

## Abstract

Wearable and lightweight devices facilitate real-world physical activity (PA) assessments. MX metrics, as a cut-point-free parameter, evaluate acceleration above which the most active X minutes are accumulated. It provides insights into the intensity of PA over specific durations. This study evaluated the association of MX metrics and clinical tests in older adults recovering from proximal femoral fracture (PFF). Analyses were conducted on the PFF cohort from the baseline assessment of the Mobilise-D project using an accelerometer-based device. Participants (N=396) were categorized into four recovery groups: acute, post-acute, extended recovery, and long-term recovery. Mobility capacity was assessed through the 6-min walking test (6MinWT), Short Physical Performance Battery (SPPB), 4-meter walking test (4MWT), and hand grip (HG) strength. Mobility perception was evaluated using the Late-Life Function and Disability Instrument (LLFDI). Eight MX metrics (M1-M90) were calculated using the GGIR package in R. Results showed moderate to strong positive correlation between M1-M30 and lower limb mobility capacity tests, and mobility perception (Lower Extremity domains) particularly in the extended and long-term recovery groups. MX metrics can be used for measuring PA intensity among older adults recovering from PFF. Shorter duration of MX metrics had higher association with lower limb mobility capacity and perception outcomes.

**Highlights:** *What are the main findings?:*

- Clinical lower limb assessments (both subjective and objective) were more discriminative in differentiating between the four PFF recovery groups in older adults.
- Older adults in the acute proximal femoral fracture recovery group demonstrated lower physical activity intensity compared to those in later recovery groups, with the differences being more pronounced for shorter-duration MX metrics (M1-M5)

*What is the implication of the main finding?:*

- The cut-point free method (e.g., MX metrics) is useful for measuring physical activity magnitude of older adult recovering from proximal femoral fracture.
- Higher lower limb capacity and perception outcomes were strongly correlated with greater daily activity intensity, particularly in older adults at later stages of proximal femoral fracture recovery.

## 1. Introduction

Physical activity (PA) is essential for various health outcomes and maintaining independence at older ages [1]. After hip fracture, individuals experience a sudden decline in their PA levels [2]. The primary objective of rehabilitation after surgery is to restore mobility and reduce disability, mortality rate, and healthcare burden [3]. As part of established clinical routines, clinicians assess patients’ PA to evaluate the effectiveness of therapeutic interventions and rehabilitation programs, and to address patients’ specific needs.

Conventionally, clinicians assess patients’ mobility capacity using common clinical tests such as the Short Physical Performance Battery (SPPB), 4-meter walking test (4MWT), 6-min walking test (6MinWT) and hand grip (HG) dynamometer [3,4]. Additionally, the Late-Life Function and Disability Instrument (LLFDI) is a widely used questionnaire for assessing mobility perception in relation to function and disability [5,6]. However, these measurements can be affected by recall bias, ceiling or floor effects, and the Hawthorne effect, particularly in controlled settings [7,8].

Wearable digital devices, such as miniaturized accelerometers, facilitate the continuous capture of real-world mobility performance (daily PA) particularly in large scale cohorts and overcome the above-mentioned limitations [9,10]. Triaxial accelerometers measure the body’s acceleration along three axes as a proxy of PA intensity and duration [10]. The traditional approach to analyzing PA intensity from accelerometer data is the cut-point method. This method relies on predefined absolute intensity cut-points to categorize the time spent in various levels of intensity achieved during various physical activities (for example, time spent in sedentary, light, and moderate-to-vigorous PA (MVPA)) [11,12]. However, this method has multiple limitations. First, intensity cutpoints are protocol- and population-dependent [9,13,14]. Therefore, it is challenging to compare or pool datasets. Second, it could easily classify PA if it has a score just below or above the cut-point [11,15]. Third, many individuals fail to achieve any activity above established cut-points. For example, prevalence of meeting guidelines (60 min/d of MVPA for children) varied from 8% to 96% depending on cut-points and sensor location [16]. Hence, there has been a recent shift from cut-point based metrics to raw acceleration data-driven metrics which are free from predefined cut-points [13,17,18].

Cut-point free metrics such as the MX metric identify the minimum acceleration value (measured in milligravitational units (mg)) above which the most active number of X minutes are accumulated during a monitoring period [11,18]. Active minutes in this metric can be accumulated across the day, which aligns with physical activity guidelines [19]. For example, if M10 of a person equals 55 mg, it means that the minimum acceleration for that person’s most active 10 minutes over 24 hours was 55 mg. This method allows for the comparison of acceleration data to any cut-point (e.g., 55 mg equivalent for MVPA level among older adults) or to an acceleration that is indicative of a standard activity (e.g., 250 mg equivalent for brisk walking among adults) [15,18,20]. Although MX metrics are population independent, their value can vary depending on age, sex, health status, sensor placement, and so on. For instance, the mean value of M10 for 12-14 years old boys’ and girls’ students was 634.4 mg and 417.1 mg respectively, who wore an accelerometer for up to 7 consecutive days on the non-dominant wrist [17]. In contrast, the M10 values measured with the same sensor placement were ∼280 mg for office workers (mean age: 44.7 years) and ∼200 mg for individuals with chronic disease (mean age: 65.2 years) [17].

Currently, research involving the MX metric has examined its association with health indicators (for example, body mass index (BMI), waist-to-height ratio, cardiorespiratory fitness) and its ability to thoroughly profile and compare physical activity intensity across different populations (such as primary school students, centenarians, pre- and post-menopausal women) [11,15,21]. However, there is a lack of knowledge regarding the relationship between MX metrics and clinical tests in different populations with health conditions such as older adult recovering from Proximal Femoral Fracture (PFF).

The main purpose of this study was to investigate the association of the MX metrics and various clinical tests including LLFDI outcomes, 6MinWT, 4MWT, SPPB, and HG strength. To this end, we analyzed the PFF cohort from the baseline assessment (T1) of the clinical validation study (CVS) of the Mobilise-D project [22]. Before measuring correlation, we classified our sample into four recovery groups that are formed based on the number of days between surgery and T1. Accordingly, we hypothesized that (1) clinical assessments and MX metrics would differ significantly across different PFF recovery groups; (2) the association between different duration of MX metrics and clinical tests would vary depending on the recovery group.

## 2. Materials and Methods

### 2.1. Design

This is a cross-sectional study using baseline data of the PFF cohort as part of Mobilise-D CVS project (Clinical Trial Registry Number: ISRCTN12051706).

### 2.2. Participants

Among 513 participants, 399 participants wore the AX6 device (Axivity, York, UK) and 114 wore the DynaPort MM+ device (McRoberts, The Hague, The Netherlands). Due to different device attachment method (directly via adhesive patch vs indirectly fixed on the belt), this study included only the participants who wore the AX6 device. Of the 399 participants, three of them were excluded due to lack of valid recorded days (see below). Thus, in total number of participants from whom we used the data was 396 (257 females, 139 males). The full inclusion and exclusion criteria are described elsewhere [22]. All participants provided written informed consent prior to data collection. Ethical approvals were obtained from Committee of the Protection of Persons, South-Mediterranean II, Montpellier (ref.: 221BO8), the ethics committee of the Medical Faculty of Eberhard-Karls-University Tubingen, Stuttgart (ref.: 976/2020BO2), the ethics committee of the Medical Faculty at Heidelberg University (ref.: S-719/2021), and the Regional Committee for Medical and Health Professional Research Ethics, Trondheim (ref.: 216069).

Participants were classified into four groups based on number of days between surgery date and clinical assessment date. Acute group: days ≤ 14, post-acute group: 14 < days ≤ 42, extended recovery group: 42 < days ≤ 182, long-term recovery group: days > 182.

### 2.3. Tasks and Procedures

#### 2.3.1. Clinical setting

Clinical outcome assessments

Mobility capacity of the participants was evaluated using SPPB, 4MWT, 6MinWT and HG strength [4,22]. The SPPB and 6MinWT were performed in a straight, hard-surface, and flat corridor. The SPPB consists of three components including static balance, a five-times chairrise test, and 4MWT [23]. Each component is scored between 0-4, and the total SPPB score spans from 0 (worst) to 12 (best). The 4MWT was repeated twice in a straight line at a comfortable and self-selected pace. The fastest trial was recorded as the maximum self-selected walking speed during the 4MWT. For 6MinWT, experimenters instructed participants to walk as far as possible within six minutes, moving back and forth along a 20-meter corridor between two cones.. Note, acute group did not perform 6MinWT due to the long duration of the test. HG strength (kg) was measured using hand dynamometer. The highest score of three attempts at both sides was used in this study.

##### Patient-reported outcome measures

The LLFDI contains two main components: Function and Disability [24]. The Disability component of LLFDI describes the *Frequency* of participating in life activities and *Limitations* in the capability of participating in those life activities. The Frequency is classified into Social and Personal Roles domains. The Limitation is classified in Instrumental Role and Management Role domains. The Function component of the LLFDI assesses task difficulty. LLFDI-Function is divided into 3 domains, Upper Extremity, Basic Lower Extremity, and Advanced Lower Extremity [24]. All items of the LLFDI components are scored on a five-point scale. The raw LLFDI scores are transformed into a scale ranging from 0-100 for easy clinical interpretation. Higher scores mean better performance and less limitation [24]. Note, LLFDI answers of patients in the acute group were related to their perception prior to the femoral fracture (pre-fracture).

#### 2.3.2. Daily life setting

To assess mobility performance, participants wore a single wearable device (AX6) which was attached directly on the lower back using a custom designed adhesive patch. Device was a 6 degrees of freedom inertial measurement unit with the following configuration; triaxial accelerometer with a range of ±8 g and resolution 1mg, triaxial gyroscope with a range of ±2000 degrees per second (dps) and a resolution of 70 mili-dps, sampling frequency 100 Hz). Participants were asked to keep the device on their lower back for 24 hours/day over 7 consecutive days. The device’s battery life allows for one week of recording without recharging. In this study, we only use triaxial accelerometer data.

#### 2.3.3. Accelerometer processing

The raw .csv data was processed using the GGIR package (version 3.1-4) of the statistical programming language R (version 4.3.1). Signal processing included 1) autocalibration using local gravity as a reference, 2) non-wear time detection, and 3) calculation of dynamic acceleration corrected for gravity (Euclidean Norm minus 1g with negative values rounded up to zero, ENMO) averaged over 5s epochs and expressed in mg units (1mg = 0.00981 m.s^-2^). Nonwear was estimated based on standard deviation and value range of each axis, calculated in 60 minutes windows with 15-minute sliding windows. Non-wear time was detected if the standard deviation for at least two out of three axes was less than 3 mg or if the value range for at least two out of three axes was less than 50 mg (Hees algorithm: F1 performance of 0.88) [25,26]. Participants were excluded if they had less than 3 valid days (defined as >14 h per day) [27]. Table A (supplementary material, available online) provides the GGIR configuration for the reproducibility principle.

Finally, eight different MX metrics (M1, M2, M5, M10, M15, M30, M60, M90 (mg)) were calculated and averaged across all valid days and wear time to provide a comprehensive picture of physical activity [17]. MX metrics were extracted in part 2 of the GGIR package using the “qlevels” argument (e.g. for M90 qlevel = (1440 - 90)/1440). These MX statistics rank the acceleration for each epoch during the day in descending order to obtain the acceleration above which the person’s most active X minutes are accumulated [15,28]. Table B of Supplementary Material (available online) provides the variable names from the GGIR output.

#### 2.3.4. Statistical Analysis

Kolmogorov–Smirnov tests revealed that all variables were not normally distributed (*p* < 0.05). Thus median, quartiles range (P25-P75), and range [minimum-maximum] were used as non-parametric descriptive statistics. One-way non-parametric ANOVA (Kruskal-Wallis with Dunn’s post hoc tests) was used to compare the four recovery groups regarding participant characteristics, clinical tests, LLFDI domains, and MX metrics. Spearman Rank correlation was conducted to assess association of MX metrics, clinical tests. The strength of the correlation value (*r*_*s*_) was classified as very weak (*r*_*s*_: 0.00-0.19), weak (*r*_*s*_: 0.20-0.39), moderate (*r*_*s*_: 0.40-0.59), strong (*r*_*s*_: 0.60-0.79), and very strong (*r*_*s*_: 0.80-1.00) [29].

A significant difference was set at a *p* level < 0.05; Bonferroni correction was applied to account for multiple testing. All statistical analyses were conducted using SPSS (version 29.0.1; Armonk, NY) and RStudio (version 2024.04.2, Boston, MA). Visualisation was done in Matlab (version R2023a; Mathworks, Natick, MA, USA)

## 3. Results

Participant characteristics are displayed in Table 1. There were three patients with missing surgery date. Therefore, these participants were not classified into recovery groups. The median (P25-P75) age of all participants was 79 (71-83) years. Post-acute participants were 5 years older than participants in the extended recovery group (*p*=0.007). The overall BMI of the participants was 23.9 (21.5-26.5) kg/m2 and was similar across the four recovery groups.

**Table 1.**
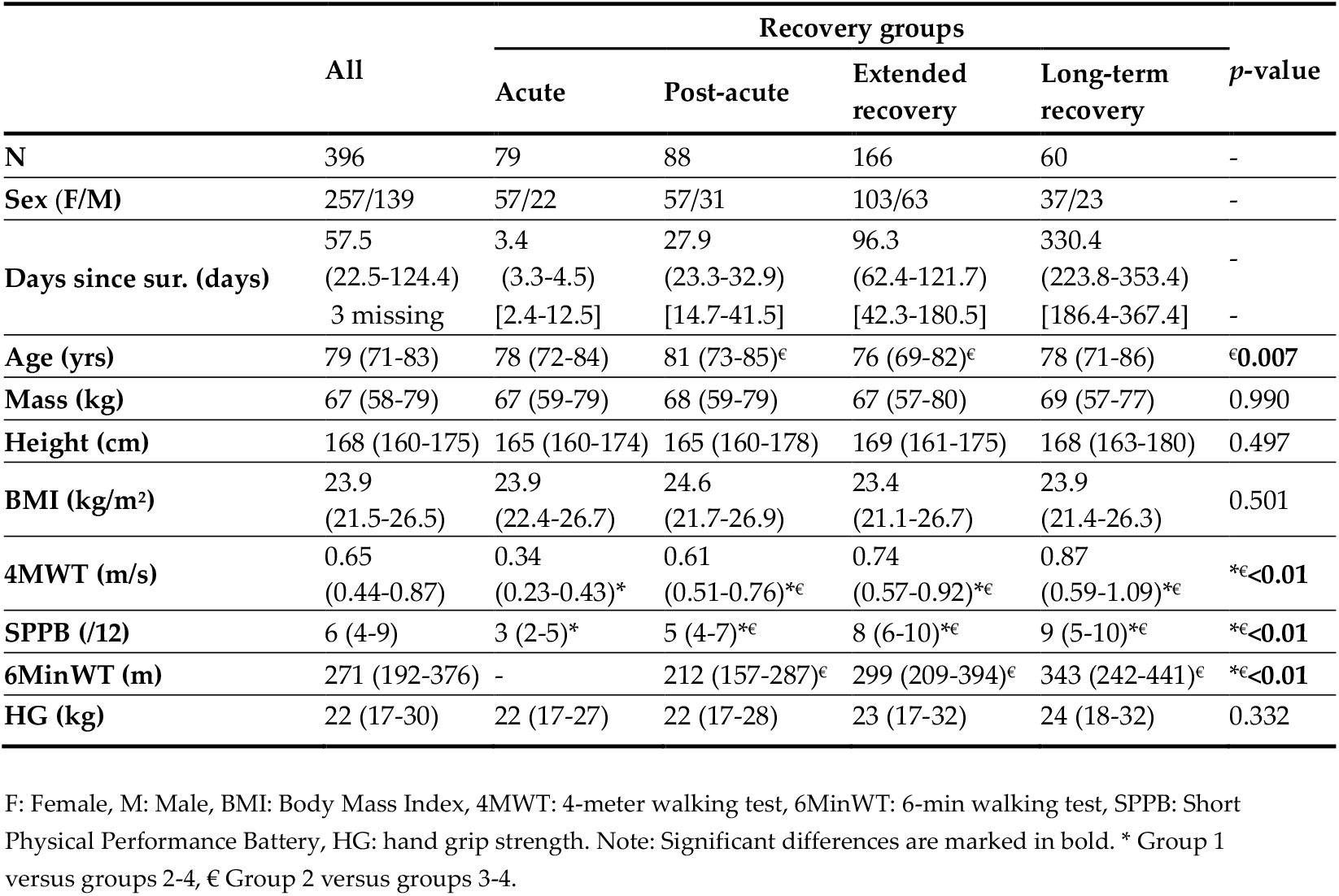
Participant characteristics and clinical tests (Median (P25-P75), [min-max]).

| | All | Recovery groups | | | | $p$-value |
| --- | --- | --- | --- | --- | --- | --- |
|  |  | Acute | Post-acute | Extended recovery | Long-term recovery |  |
| N | 396 | 79 | 88 | 166 | 60 | - |
| Sex (F/M) | 257/139 | 57/22 | 57/31 | 103/63 | 37/23 | - |
| Days since sur. (days) | 57.5<br>(22.5-124.4) | 3.4<br>(3.3-4.5) | 27.9<br>(23.3-32.9) | 96.3<br>(62.4-121.7) | 330.4<br>(223.8-353.4) | - |
|  | 3 missing | [2.4-12.5] | [14.7-41.5] | [42.3-180.5] | [186.4-367.4] | - |
| Age (yrs) | 79 (71-83) | 78 (72-84) | 81 (73-85)<sup>€</sup> | 76 (69-82)<sup>€</sup> | 78 (71-86) | <sup>€</sup>**0.007** |
| Mass (kg) | 67 (58-79) | 67 (59-79) | 68 (59-79) | 67 (57-80) | 69 (57-77) | 0.990 |
| Height (cm) | 168 (160-175) | 165 (160-174) | 165 (160-178) | 169 (161-175) | 168 (163-180) | 0.497 |
| BMI (kg/m<sup>2</sup>) | 23.9<br>(21.5-26.5) | 23.9<br>(22.4-26.7) | 24.6<br>(21.7-26.9) | 23.4<br>(21.1-26.7) | 23.9<br>(21.4-26.3) | 0.501 |
| 4MWT (m/s) | 0.65<br>(0.44-0.87) | 0.34<br>(0.23-0.43)* | 0.61<br>(0.51-0.76)<sup>*€</sup> | 0.74<br>(0.57-0.92)<sup>*€</sup> | 0.87<br>(0.59-1.09)<sup>*€</sup> | <sup>*€</sup>**<0.01** |
| SPPB (/12) | 6 (4-9) | 3 (2-5)* | 5 (4-7)<sup>*€</sup> | 8 (6-10)<sup>*€</sup> | 9 (5-10)<sup>*€</sup> | <sup>*€</sup>**<0.01** |
| 6MinWT (m) | 271 (192-376) | - | 212 (157-287)<sup>€</sup> | 299 (209-394)<sup>€</sup> | 343 (242-441)<sup>€</sup> | <sup>*€</sup>**<0.01** |
| HG (kg) | 22 (17-30) | 22 (17-27) | 22 (17-28) | 23 (17-32) | 24 (18-32) | 0.332 |

F: Female, M: Male, BMI: Body Mass Index, 4MWT: 4-meter walking test, 6MinWT: 6-min walking test, SPPB: Short Physical Performance Battery, HG: hand grip strength. Note: Significant differences are marked in bold. \* Group 1 versus groups 2-4, € Group 2 versus groups 3-4.

The median value of mobility capacity outcomes is depicted in a radar plot (Figure 1). The four axes displayed in this plot are 4MWT, SPPB, 6MinWT and HG tests with values increasing outward from the centre. In general, 4MWT speed and SPPB score showed the most significant differences between groups. Distance covered during 6MinWT test in extended recovery and long-term recovery were 87 cm and 131 cm longer than in the post-acute group, respectively (all *p* < 0.01). The HG strength was similar among the participants in the four different recovery groups (*p* = 0.332) (Table 1, Figure. 1).

**Figure 1.**
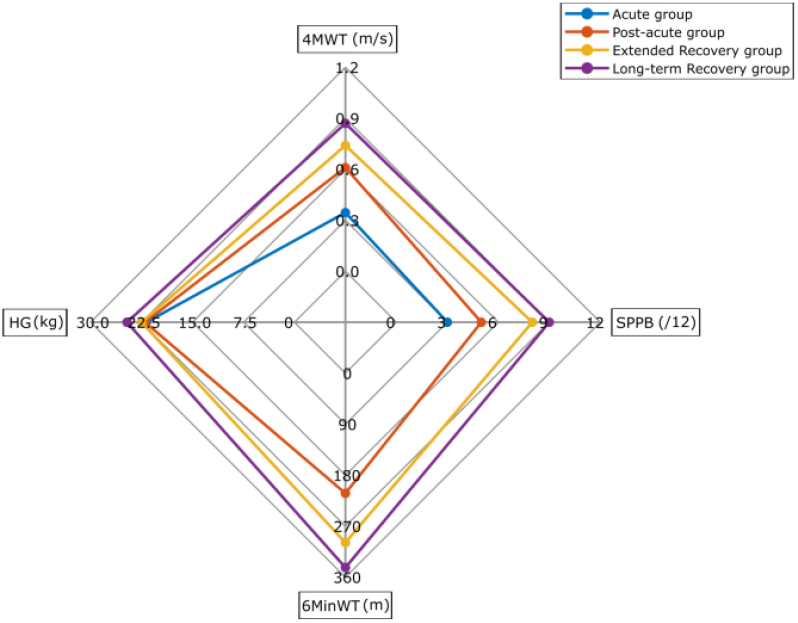
Radar plot illustrating median of 4-meter walking test (4MWT), Short Physical Performance Battery (SPPB) score, 6-min walking test distance (6MinWT), and Hand Grip (HG) strength in four recovery groups of PFF patients.

Figures 2 and 3 contain the box plots of the LLFDI components Disability and Function. The horizontal line inside the box represents the median value. Each box indicates the quartiles range. The median value of Personal Role, Social Role, and Instrumental Role scores in acute group (pre-fracture) were significantly higher than for the participants in the post-acute and extended recovery groups (all *p* < 0.05). In addition, the median value of the Management Role score of participants in the post-acute group was lower than in the acute group (pre-fracture) and extended recovery groups (all *p* < 0.05). The Personal Role, Social Role, and Instrumental Role scores among participants in extended and long-term recovery groups were higher than in the post-acute patients (all *p* < 0.05) (Figure 2). Appendix C in the supplementary material displays the median (P25–P75) values and between group comparisons for all LLFDI domains.

**Figure 2.**
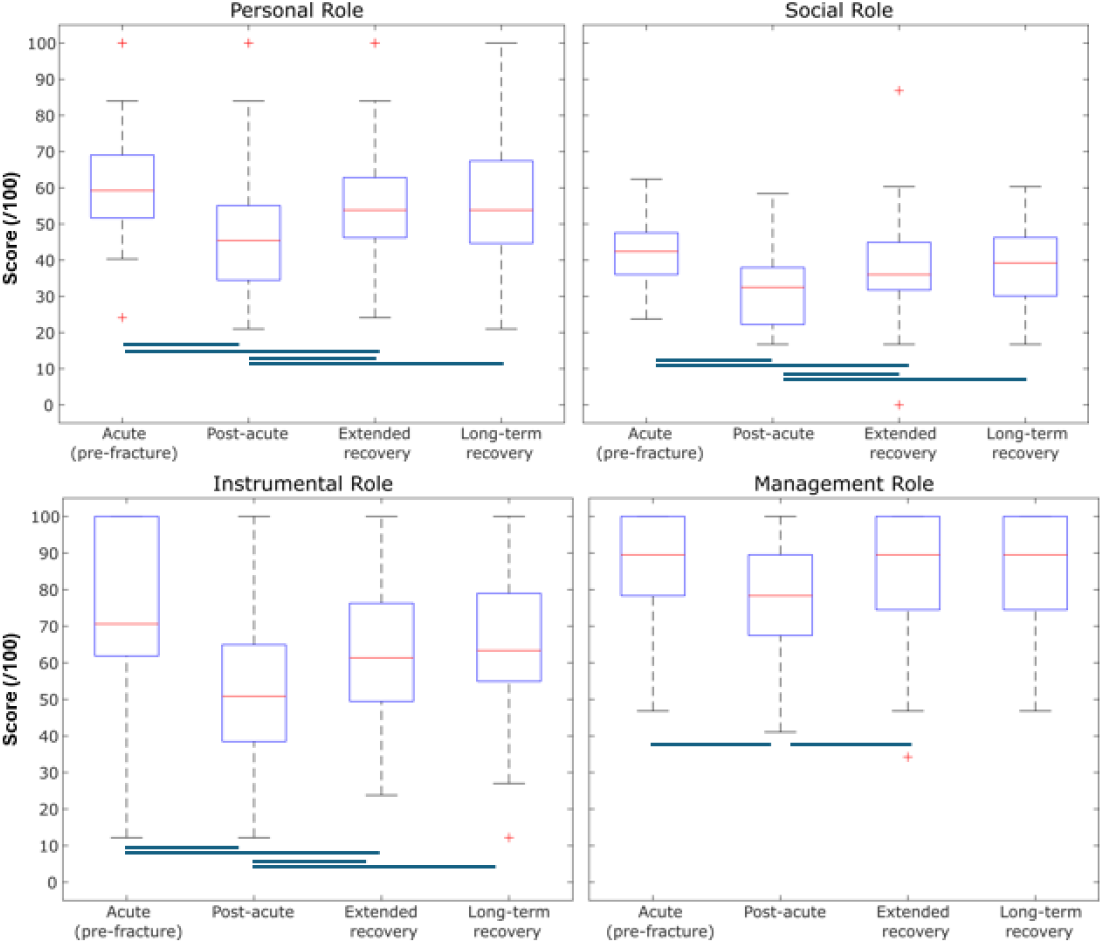
Box plot illustrating the Disability component of the LLFDI among four PFF recovery groups (whiskers extend to data points within 1.5 times the interquartile range, outliers marked as red plus, blue lines mean significant difference (*p* < 0.05)).

**Figure 3.**
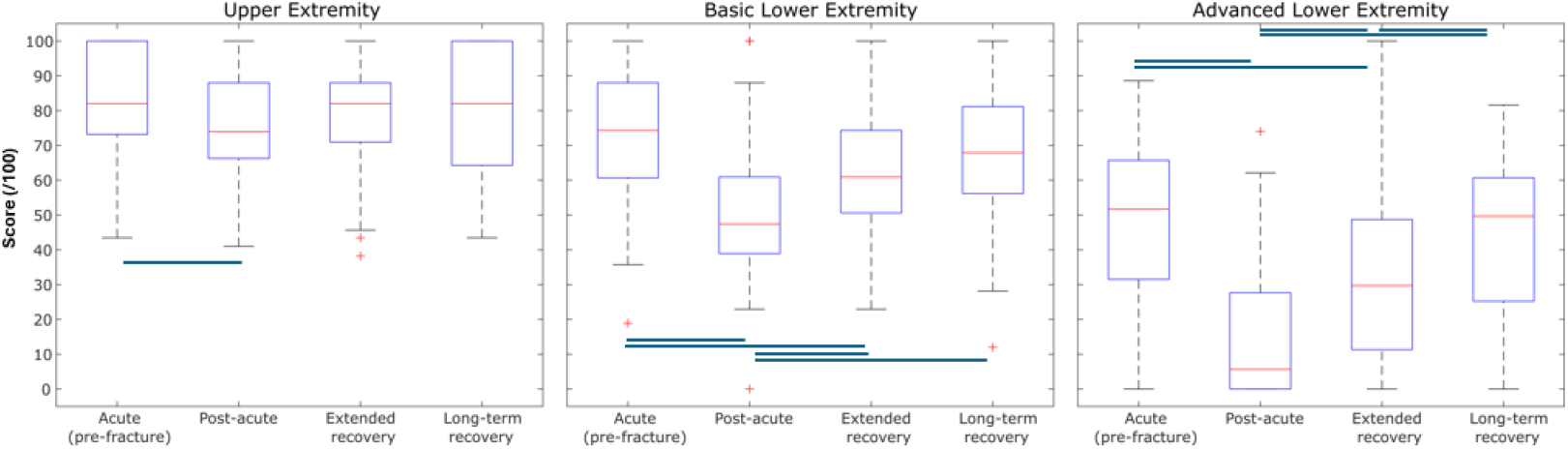
Box plot illustrating Function component of LLFDI score among four PFF recovery groups (Whiskers extend to data points within 1.5 times the interquartile range, outliers marked as red plus, blue lines mean significant difference (*p* < 0.05)).

As can be seen in Figure 3, Advanced Lower Extremity domain of LLFDI had the most between group differences. Median values of the Upper Extremity, Basic Lower Extremity, and Advanced Lower Extremity scores among acute group (pre-fracture) were higher than those in the post-acute and extended recovery groups (all *p* < 0.05). Additionally, median values of Basic Lower Extremity and Advanced Lower Extremity scores of participants in extended and long-term recovery groups were higher than those in the post-acute group (all *p* < 0.05) (Figure 3).

The violin plots in Figure 4 display the distribution of MX metrics’ intensity among the four recovery groups. Each point in the plot represents an individual value. M1, M2, and M5 had the most between group differences. The intensity of all MX metrics was significantly higher in post-acute group compared to the acute group. Similarly, extended and long-term recovery groups had a higher intensity of MX metrices compared to acute group except for M90. Apart from M1, there was no difference between post-acute and extended recovery groups. Appendix D in the supplementary material presents the median (P25–P75) values and the between group comparisons of the selected MX metrics.

**Figure 4.**
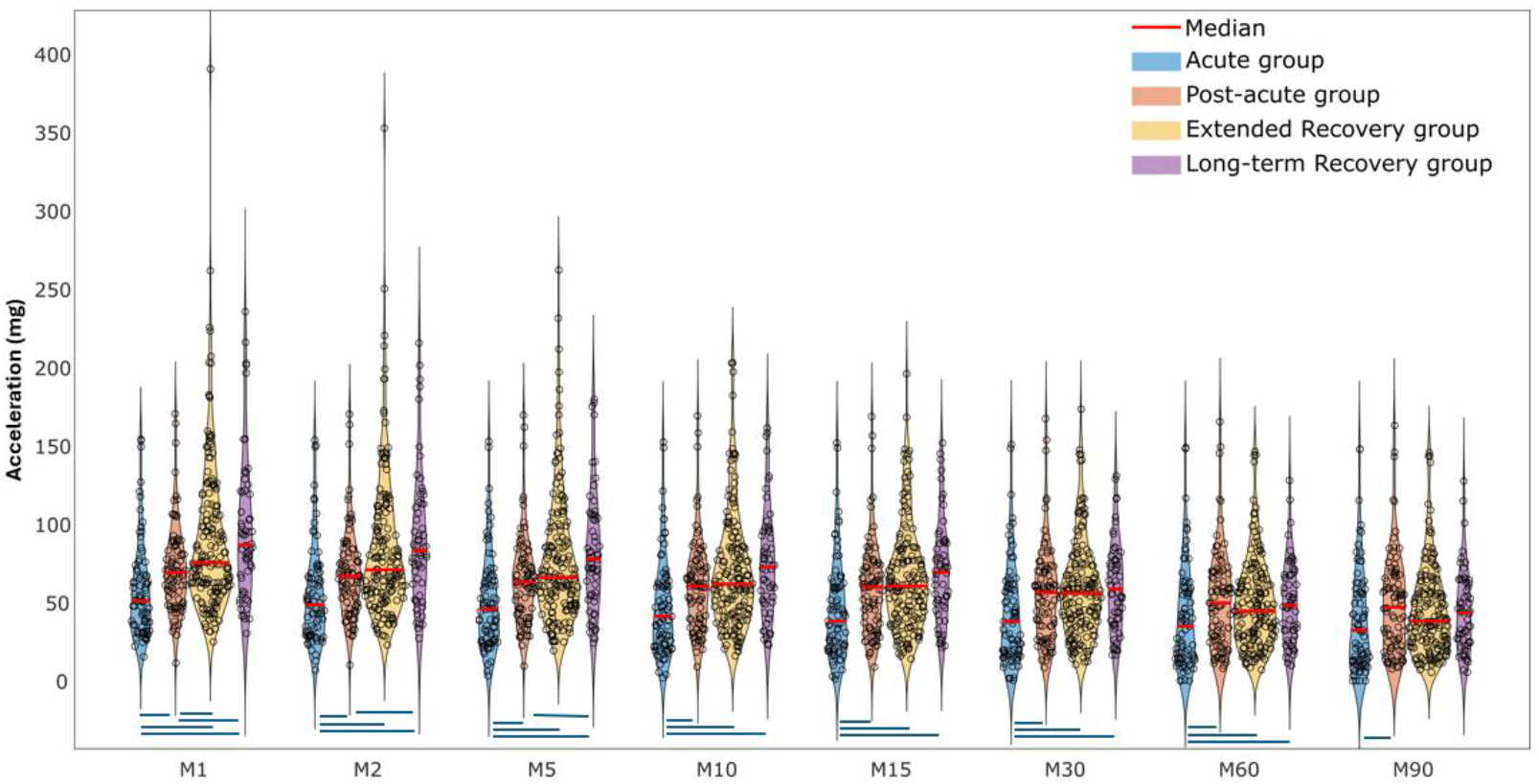
Violin plot illustrating MX metrics among four recovery groups (blue lines mean significant difference (p < 0.05)).

The heat map in Figure 5 depicts the correlations between clinical tests (y-axis: 4MWT, SPPB, 6MinWT, HG, seven LLFDI’s domains) and MX metrics (x-axis) for all participants as well as each recovery group. The colour intensity ranges from light blue (very week correlation) to dark blue (strong correlation). The darkest blue areas of clinical assessments belonged to the relationship of 6MinWT, 4MWT, SPPB and M1-M30. The darkest blue area of LLFDI domains belonged first to function component (Basic Lower Extremity and Advanced Lower Extremity domains) then to Disability component (Social Role and Instrumental Role domains) and M1-M30. Moreover, these relationships are stronger among participants in extended and long-term recovery groups, particularly with shorter MX metrics durations (M1-M10) (Figure 5).

**Figure 5.**
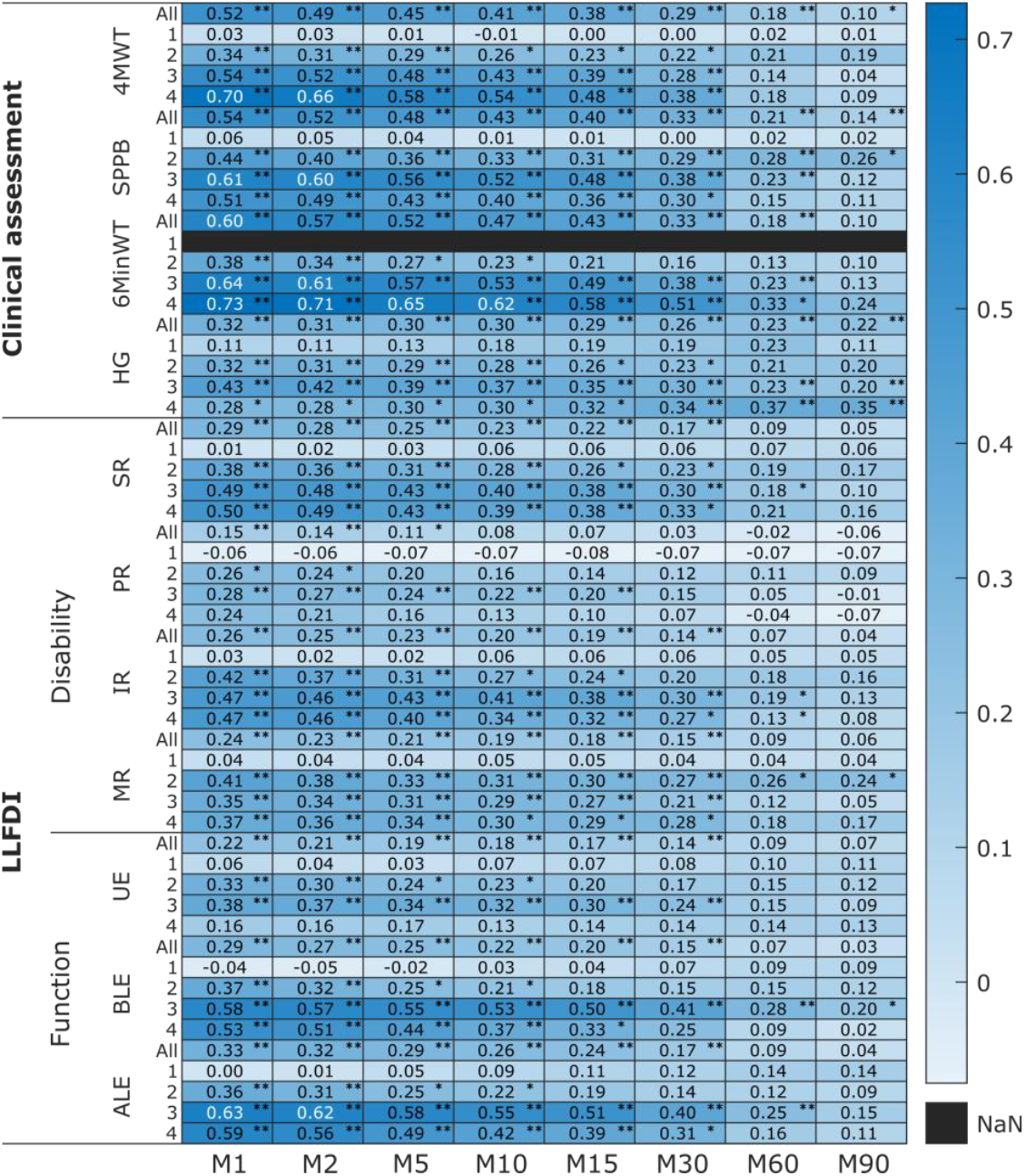
Heat map illustrating the level of association (*r*_*s*_ value) between MX metrics (M1-M90) and all clinical tests among PFF patients in four recovery groups: 1) acute (pre-fracture for LLFDI assessments), 2) post-acute, 3) extended recovery, 4) long-term recovery, All) all participants (N=396). Clinical tests: 4-meter walking test (4MWT), Short Physical Performance Battery (SPPB) score, 6-min walking test distance (6MinWT), and Hand Grip (HG) strength, LLFDI domains: Social Role (SR), Personal Role (PR), Instrumental Role (IR), and Management Role (MR), Upper Extremity (UE), Basic Lower Extremity (BLE), Advanced Lower Extremity (ALE) (*: *p* < 0.05, ** *p* < 0.01, white font colour: strong correlation (*r*_*s*_: 0.60-0.79), NaN: not a number).

## 4. Discussion

To the best of our knowledge, this is the first study to analyze the association between cut-point free metrics (MX) and clinical tests (mobility capacity and perception) among older adults recovering from PFF. To this end, we first compared MX metrics and clinical tests across the four recovery categories of our participants. Subsequently, we analyzed the relationships between MX metrics and clinical tests for all participants as well as within each recovery category.

Demographic characteristics in the four recovery groups were similar (Table 1). The mobility capacity of our participants was measured using objective clinical tests. Based on results (SPPB, 4MWT, and 6MinWT outcomes), mobility capacities such as walking speed, balance, lower limb strength, and endurance were higher among participants in later recovery groups (Figure 1). The 4MWT and SPPB revealed the most between group differences. These findings suggest that the 4MWT and SPPB have a higher potential for detecting meaningful differences among PFF patients at various recovery stages. Better mobility capacity outcomes (e.g., walking speed, balance, and endurance) in extended recovery and long-term recovery groups can reflect positive effect of rehabilitation program and longer recovery duration.

Mobility perception scores, as assessed through seven different domains of the LLFDI questionnaire. Advance Lower Extremity domain of LLFDI (involving activities that require a high level of physical ability and endurance like running 1/2 mile) has the most between group difference (Figure 3). Then, it has a potential to be responsive to meaningful between group differences. Mobility perceptions were higher among extended and long-term recovery groups compared to post-acute group which was aligned with mobility capacities findings. However, mobility perception among our participants after PFF (post-acute, extended, and long-term recovery groups) was lower than acute group before PFF (Figure 2, 3). These findings reveal that despite the improvement in mobility perception over time among our participants after PFF, it remained lower than acute group (pre-fracture). However, the mobility perception of our patients before fracture belonged to our participants in acute group which could have been affected by recall bias and pain. Our findings aligned with previous studies [30,31] which reported inactivity and intensity of pain can be considered as important risk factors for worse self-perceived health and functional mobility. Further studies are needed to evaluate longitudinal LLFDI among older adults recovering from PFF.

The violin plot visualized MX metrics as a duration-related PA intensity among older adults in four PFF recovery groups. The shorter durations of MX metrics (M1-M5) with greater intensity were more effective in distinguishing differences among our participants (Figure 4). Intensity of PA longer than 10 minutes (M10-M90) were similar across post-acute, extended, and long-term recovery groups. Our findings were aligned with Rowlands and his colleagues (2019, Fig. 1) that found bigger between group differences (adolescent, office workers, adult with type 2 diabetes) for short time periods and higher intensity PA (i.e. M5, M15). Therefore, these findings confirm our first hypothesis about difference of mobility capacity, mobility perception, and MX metrics among PFF patients in different recovery stages after surgery.

One approach to interpret MX metrics is using cut-points (e.g., sedentary, light, MVPA) [20]. Duncan and colleagues (2020) calibrated cut-points of acceleration data in older adults based on energy expenditure [20]. They identified the following cut-points for sedentary time (11.7 mg), light physical activity (11.7-54.9 mg) and moderate-to-vigorous (MVPA) (55 mg) in older adult from waist-worn accelerometers. Notably, their study protocol did not include vigorous physical activity (VPA). Median values of our MX metrics for PFF patients in acute group fell within the light intensity category and it gradually decreased from M1: 51.1 mg to M90: 32.4 mg. For PFF patients in post-acute, extended and longterm recovery groups, the median values for M1–M30 exceeded the MVPA threshold. However, for M60 and M90, the median values across all recovery groups declined and fell into the light intensity category. Therefore, MX metrics provide a clearer comparison of duration-related PA intensity and demonstrate better between-group differences, particularly when the PA levels of different groups fall within the same category.

Rowlands and his colleagues (2021) suggested another approach to interpret accelerometery data [32]. Their study was among inactive adult UK Biobank using wrist-worn accelerometers. Their research suggests that 1.0 mg increase in daily average acceleration is equal to 5-6 min brisk walking (500 steps taken in 5 minutes) which is associated with a greater life expectancy of 3.9 years and 5% decreased in all-case mortality. In this study, we did not measure daily average acceleration. However, an increase in MX metrics would lead to an increase in daily average acceleration. Therefore, MX metrics as a proxy for time-related intensity can be used for surveillance of PA magnitude among PFF patients.

For the second hypothesis of this study, we found a strong correlation between lower limb clinical assessments (4MWT, SPPB, 6MinWT), the Function component of LLFDI (Advanced Lower Extremity), and shorter durations of MX metrics (M1-M30). This suggests a higher association between mobility capacity, functional perception of lower limb activities, and shorter durations of daily PA among older adults recovering from PFF. Notably, this association was stronger among participants in the later recovery groups (extended and longterm recovery groups). This finding also indicates that participants in the later recovery stages, with higher mobility capacities, could perform daily PA at higher intensities, which could potentially contribute to improved overall health and life expectancy.

Stamatakis and his colleagues, 2022, analyzed wrist-worn accelerometer of 103,684 UK Biobank adults [33]. Their results revealed that approximately 3-4 minutes of vigorous intermittent lifestyle physical activity (VILPA) were associated with substantial lower mortality risk. They also found that VILPA in nonexercisers can have similar effects to VPA in exercisers, suggesting that VILPA (i.e. may be a suitable PA target particularly in individuals not able or willing to exercise. A longitudinal study about the mortality rate and shorter duration of MX metrics as VILPA could provide a new insight of daily PA among different population.

The association between M1-M10, Social Role, and Instrumental Role were positive and moderate among our participants. This means higher magnitude of daily PA is moderately associated with higher perception of engaging in social activities and less limitations in activities both at home and in the community among older adults recovering from PFF. These findings support our second hypothesis, which proposed varying levels of association between MX metrics and clinical measurements in PFF patients. Based on our findings we can suggest more sophisticated analysis on shorter duration of MX metrics and clinical lower limb assessments both for within and between in different populations. Then MX metrics would provide a public health-friendly and personalized interpretation of PA with the potential to be a standardized accelerometer outcome as a guideline [18,21].

The limitations of this study included the unequal sample sizes across recovery groups, sex heterogeneity, and the absence of contextual information for both clinical and daily PA assessments. Additionally, monitoring daily PA using a sensor may introduce a Hawthorne effect. The placement of the sensor on the lower back, where it is less visible, could help mitigate this effect. However, we are confident that this preliminary study provides practical insight for clinicians and researchers to apply cut-point free metrics such as MX to precisely analyze PA and fill the gap between cut-point free metrics and traditional assessment.

## 5. Conclusions

Clinical lower limb functional assessments, whether mobility capacity tests (e.g., 4MWT) or mobility perception tests (e.g., LLFDI), were more discriminative in differentiating between the four PFF recovery groups among our older adult participants. Shorter duration of MX metrics (M1-M5) with higher intensity were more effective for between group comparison. Older adults in the later recovery stages, with higher mobility capacities could perform daily PA at higher intensities, which could potentially contribute to improved overall health and life expectancy. The associations between MX metrics and clinical tests (both mobility capacity and perception assessments) were positive, and stronger in shorter duration and higher magnitude of MX, particularly among PFF patients in later recovery groups.

## Supplementary Materials

The following supporting information can be downloaded at: www.mdpi.com/xxx/s1, Figure S1: title; Table S1: title; Video S1: title.

## Author Contributions

H.Y.: Conceptualization, methodology, formal analysis, software, visualization, writing - original draft; D.S: data curation; B.V, J.G.A., L.R., M.A.B, M.E., J.B., S.K., C.P.J., K.A., A.I., and C.B.: methodology, writing - review & editing; P.A., J.L.H: methodology; B.C: conceptualization, methodology, writing - review & editing, supervision.

## Institutional Review Board Statement

The study was conducted in accordance with the Declaration of Helsinki, and approved by Committee of the Protection of Persons, South-Mediterranean II, Montpellier (ref.: 221BO8), the ethics committee of the Medical Faculty of Eberhard-Karls-University Tubingen, Stuttgart (ref.: 976/2020BO2), the ethics committee of the Medical Faculty at Heidelberg University (ref.: S-719/2021), and the Regional Committee for Medical and Health Professional Research Ethics, Trondheim (ref.: 216069).

## Informed Consent Statement

Informed consent was obtained from all subjects involved in the study. Written informed consent has been obtained from the patient(s) to publish this paper” if applicable.

## Data Availability Statement

The data in this study is not currently available outside the Mobilise-D consortium. However, the Mobilise-D consortium is currently planning a public data release for late 2025, which will be available on the Mobilise-D Zenodo page: https://zenodo.org/communities/mobilise-d.

## Acknowledgments

The authors would like to thank the participants who participated in the study for their time and contributions.

## Abbreviations

The following abbreviations are used in this manuscript:

PFF: proximal femoral fracture
PA: physical activity
SPPB: short physical performance battery
4MWT: 4-meter walking test
6MinWT: 6-min walking test
HG: hand grip
LLFDI: late-life function and disability Instrument
MVPA: moderate-to-vigorous physical activity
VILPA: vigorous intermittent lifestyle physical activity
BMI: body mass index
T1: baseline assessment
CVS: clinical validation study
mg: milligravitational
dps: degrees per second

## References

1. Bull, F.C.; Al-Ansari, S.S.; Biddle, S.; Borodulin, K.; Buman, M.P.; Cardon, G.; Carty, C.; Chaput, J.-P.; Chastin, S.; Chou, R. World Health Organization 2020 guidelines on physical activity and sedentary behaviour. British journal of sports medicine 2020, 54, 1451–1462, doi:10.1136/bjsports-2020-102955.

2. Zusman, E.Z.; Dawes, M.G.; Edwards, N.; Ashe, M.C. A systematic review of evidence for older adults’ sedentary behavior and physical activity after hip fracture. Clinical rehabilitation 2018, 32, 679–691, doi:10.1177/026921551774166.

3. Taraldsen, K.; Polhemus, A.; Engdal, M.; Jansen, C.-P.; Becker, C.; Brenner, N.; Blain, H.; Johnsen, L.; Vereijken, B. Evaluation of mobility recovery after hip fracture: a scoping review of randomized controlled studies. Osteoporosis International 2024, 35, 203–215, doi:10.1007/s00198-023-06922-4.

4. Xu, B.Y.; Yan, S.; Low, L.L.; Vasanwala, F.F.; Low, S.G. Predictors of poor functional outcomes and mortality in patients with hip fracture: a systematic review. BMC musculoskeletal disorders 2019, 20, 1–9, doi:10.1186/s12891-019-2950-0.

5. Haley, S.M.; Jette, A.M.; Coster, W.J.; Kooyoomjian, J.T.; Levenson, S.; Heeren, T.; Ashba, J. Late life function and disability instrument: II. Development and evaluation of the function component. The Journals of Gerontology Series A: Biological Sciences and Medical Sciences 2002, 57, M217–M222, doi:10.1093/gerona/57.4.M217.

6. Beauchamp, M.; Hao, Q.; Kuspinar, A.; Alder, G.; Makino, K.; Nouredanesh, M.; Zhao, Y.; Mikton, C.; Thiyagarajan, J.A.; Diaz, T. Measures of perceived mobility ability in community-dwelling older adults: a systematic review of psychometric properties. Age and Ageing 2023, 52, iv100–iv111, doi:10.1093/ageing/afad124.

7. McCambridge, J.; Witton, J.; Elbourne, D.R. Systematic review of the Hawthorne effect: new concepts are needed to study research participation effects. Journal of clinical epidemiology 2014, 67, 267–277, doi:10.1016/j.jclinepi.2013.08.015.

8. Stull, D.E.; Leidy, N.K.; Parasuraman, B.; Chassany, O. Optimal recall periods for patient-reported outcomes: challenges and potential solutions. Current medical research and opinion 2009, 25, 929–942, doi:10.1185/03007990902774765.

9. Troiano, R.P.; McClain, J.J.; Brychta, R.J.; Chen, K.Y. Evolution of accelerometer methods for physical activity research. British journal of sports medicine 2014, 48, 1019–1023, doi:10.1136/bjsports-2014-093546.

10. Lee, I.-M.; Shiroma, E.J. Using accelerometers to measure physical activity in large-scale epidemiological studies: issues and challenges. British journal of sports medicine 2014, 48, 197–201, doi:10.1136/bjsports-2013-093154.

11. Hernández-Vicente, A.; Marín-Puyalto, J.; Pueyo, E.; Vicente-Rodríguez, G.; Garatachea, N. Physical activity in centenarians beyond cut-point-based accelerometer metrics. International Journal of Environmental Research and Public Health 2022, 19, 11384, doi:10.3390/ijerph191811384.

12. Migueles, J.H.; Cadenas-Sanchez, C.; Alcantara, J.M.; Leal-Martín, J.; Mañas, A.; Ara, I.; Glynn, N.W.; Shiroma, E.J. Calibration and cross-validation of accelerometer cut-points to classify sedentary time and physical activity from hip and non-dominant and dominant wrists in older adults. Sensors 2021, 21, 3326, doi:10.3390/s21103326.

13. Troiano, R.P. Evolution of public health physical activity applications of accelerometers: A personal perspective. Journal for the Measurement of Physical Behaviour 2023, 6, 13–18, doi:10.1123/jmpb.2022-0038.

14. Buchan, D.S.; Maylor, B.D. Comparison of physical activity metrics from two research-grade accelerometers worn on the non-dominant wrist and thigh in children. Journal of Sports Sciences 2023, 41, 80–88, doi:10.1080/02640414.2023.2197726.

15. Rowlands, A.V.; Sherar, L.B.; Fairclough, S.J.; Yates, T.; Edwardson, C.L.; Harrington, D.M.; Davies, M.J.; Munir, F.; Khunti, K.; Stiles, V.H. A data-driven, meaningful, easy to interpret, standardised accelerometer outcome variable for global surveillance. Journal of Science and Medicine in Sport 2019, 22, 1132–1138, doi:10.1016/j.jsams.2019.06.016.

16. Migueles, J.H.; Cadenas-Sanchez, C.; Tudor-Locke, C.; Löf, M.; Esteban-Cornejo, I.; Molina-Garcia, P.; Mora-Gonzalez, J.; Rodriguez-Ayllon, M.; Garcia-Marmol, E.; Ekelund, U. Comparability of published cut-points for the assessment of physical activity: Implications for data harmonization. Scandinavian journal of medicine & science in sports 2019, 29, 566–574, doi:10.1111/sms.13356.

17. Fairclough, S.J.; Rowlands, A.V.; del Pozo Cruz, B.; Crotti, M.; Foweather, L.; Graves, L.E.; Hurter, L.; Jones, O.; MacDonald, M.; McCann, D.A. Reference values for wrist-worn accelerometer physical activity metrics in England children and adolescents. International Journal of Behavioral Nutrition and Physical Activity 2023, 20, 35, doi:10.1186/s12966-023-01435-z.

18. Rowlands, A.V.; Dawkins, N.P.; Maylor, B.; Edwardson, C.L.; Fairclough, S.J.; Davies, M.J.; Harrington, D.M.; Khunti, K.; Yates, T. Enhancing the value of accelerometer-assessed physical activity: meaningful visual comparisons of data-driven translational accelerometer metrics. Sports medicine-open 2019, 5, 47, doi:10.1186/s40798-019-0225-9.

19. Olson, R.D.; Vaux-Bjerke, A.; Quam, J.B.; Piercy, K.L.; Troiano, R.P.; George, S.M.; Sprow, K.; Ballard, R.M.; Fulton, J.E.; Galuska, D.A. Physical activity guidelines for Americans. Physical activity guidelines for Americans. NADAR! SWIMMING MAGAZINE-Periódico científico em esportes e fitness aquático-natação, pólo aquático, nado sincronizado, saltos ornamentais, travessias aquáticas. 2023.

20. Duncan, M.J.; Rowlands, A.; Lawson, C.; Leddington Wright, S.; Hill, M.; Morris, M.; Eyre, E.; Tallis, J. Using accelerometry to classify physical activity intensity in older adults: What is the optimal wear-site? European journal of sport science 2020, 20, 1131–1139, doi:10.1080/17461391.2019.1694078.

21. Fairclough, S.J.; Rowlands, A.V.; Taylor, S.; Boddy, L.M. Cut -point-free accelerometer metrics to assess children’s physical activity: An example using the school day. Scandinavian Journal of Medicine & Science in Sports 2020, 30, 117–125, doi:10.1111/sms.13565.

22. Mikolaizak, A.S.; Rochester, L.; Maetzler, W.; Sharrack, B.; Demeyer, H.; Mazzà, C.; Caulfield, B.; Garcia-Aymerich, J.; Vereijken, B.; Arnera, V. Connecting real-world digital mobility assessment to clinical outcomes for regulatory and clinical endorsement–the Mobilise-D study protocol. PLoS One 2022, 17, e0269615, doi:10.1371/journal.pone.0269615.

23. Guralnik, J.M.; Simonsick, E.M.; Ferrucci, L.; Glynn, R.J.; Berkman, L.F.; Blazer, D.G.; Scherr, P.A.; Wallace, R.B. A short physical performance battery assessing lower extremity function: association with self-reported disability and prediction of mortality and nursing home admission. Journal of gerontology 1994, 49, M85–M94.

24. Jette, A.M.; Haley, S.M.; Kooyoomjian, J.T. Late-life FDI manual. Boston, MA: Boston University 2002, 73.

25. van Hees, V.T.; Renström, F.; Wright, A.; Gradmark, A.; Catt, M.; Chen, K.Y.; Löf, M.; Bluck, L.; Pomeroy, J.; Wareham, N.J. Estimation of daily energy expenditure in pregnant and non-pregnant women using a wrist-worn tri-axial accelerometer. PloS one 2011, 6, e22922, doi:10.1371/journal.pone.0022922.

26. Syed, S.; Morseth, B.; Hopstock, L.A.; Horsch, A. Evaluating the performance of raw and epoch non-wear algorithms using multiple accelerometers and electrocardiogram recordings. Scientific Reports 2020, 10, 5866, doi:10.1038/s41598-020-62821-2.

27. Herrmann, S.D.; Barreira, T.V.; Kang, M.; Ainsworth, B.E. Impact of accelerometer wear time on physical activity data: a NHANES semisimulation data approach. British journal of sports medicine 2014, 48, 278–282, doi:10.1136/bjsports-2012-091410.

28. Rowlands, A.V.; Edwardson, C.L.; Dawkins, N.P.; Maylor, B.D.; Metcalf, K.M.; Janz, K.F. Physical activity for bone health: how much and/or how hard? Medicine & Science in Sports & Exercise 2020, 52, 2331–2341, doi:10.1249/MSS.0000000000002380.

29. Evans, J.D. Straightforward statistics for the behavioral sciences; Thomson Brooks/Cole Publishing Co: 1996.

30. Denche-Zamorano, Á.; Salas-Gómez, D.; Barrios-Fernandez, S.; Tomás-Carus, P.; Adsuar, J.C.; Parraca, J.A. Relationship Between Frequency of Physical Activity, Functional Mobility, and Self-Perceived Health in People with Different Levels of Pain: A Cross-Sectional Study. Journal of Functional Morphology and Kinesiology 2024, 9, 198, doi:10.3390/jfmk9040198.

31. Pereira, L.V.; Vasconcelos, P.P.d.; Souza, L.A.F.; Pereira, G.d.A.; Nakatani, A.Y.K.; Bachion, M.M. Prevalence and intensity of chronic pain and self-perceived health among elderly people: a population-based study. Revista latino-americana de enfermagem 2014, 22, 662–669, doi:10.1590/0104-1169.3591.2465.

32. Rowlands, A.; Davies, M.; Dempsey, P.; Edwardson, C.; Razieh, C.; Yates, T. Wrist-worn accelerometers: recommending∼ 1.0 mg as the minimum clinically important difference (MCID) in daily average acceleration for inactive adults. British journal of sports medicine 2021, 55, 814–815, doi:10.1136/bjsports-2020-102293.

33. Stamatakis, E.; Ahmadi, M.N.; Gill, J.M.; Thøgersen-Ntoumani, C.; Gibala, M.J.; Doherty, A.; Hamer, M. Association of wearable device-measured vigorous intermittent lifestyle physical activity with mortality. Nature Medicine 2022, 28, 2521–2529, doi:10.1038/s41591-022-02100-x.

