## Supplementary material for "Association of Cut-point Free Metrics and Common Clinical Tests among Older Adults after Proximal Femoral Fracture": Appendix.docx

**Appendix A.** GGIR configuration.

| **Argument** | **Value** | **Context** |
| --- | --- | --- |
| do.report | 2 | not applicable |
| f0 | 1 | not applicable |
| f1 | 396 | not applicable |
| mode | 1:2 | not applicable |
| studyname | c() | not applicable |
| GGIRread_version | 1.0.2 | not applicable |
| GGIRversion | 3.1.4 | not applicable |
| R_version | R version 4.3.1 (2023-06-16 ucrt) | not applicable |
| qwindow | c(0,24) | params_247 |
| qlevels | c(0.833333333333333,0.875,0.916666666666667,0.9375,0.958333333333333,0.965277777777778,0.972222222222222,0.979166666666667,0.986111111111111,0.989583333333333,0.993055555555556,0.996527777777778,0.998611111111111,0.999305555555556) | params_247 |
| qwindow_dateformat | %d-%m-%Y | params_247 |
| ilevels | c() | params_247 |
| IVIS_windowsize_minutes | 60 | params_247 |
| IVIS_epochsize_seconds | c() | params_247 |
| IVIS.activity.metric | 1 | params_247 |
| IVIS_acc_threshold | 5 | params_247 |
| qM5L5 | 0.25 | params_247 |
| MX.ig.min.dur | 1 | params_247 |
| M5L5res | 10 | params_247 |
| winhr | 5 | params_247 |
| iglevels | c(0,25,50,75,100,125,150,175,200,225,250,275,300,325,350,375,400,425,450,475,500,525,550,575,600,625,650,675,700,725,750,775,800,825,850,875,900,925,950,975,1000,1025,1050,1075,1100,1125,1150,1175,1200,1225,1250,1275,1300,1325,1350,1375,1400,1425,1450,1475,1500,1525,1550,1575,1600,1625,1650,1675,1700,1725,1750,1775,1800,1825,1850,1875,1900,1925,1950,1975,2000,2025,2050,2075,2100,2125,2150,2175,2200,2225,2250,2275,2300,2325,2350,2375,2400,2425,2450,2475,2500,2525,2550,2575,2600,2625,2650,2675,2700,2725,2750,2775,2800,2825,2850,2875,2900,2925,2950,2975,3000,3025,3050,3075,3100,3125,3150,3175,3200,3225,3250,3275,3300,3325,3350,3375,3400,3425,3450,3475,3500,3525,3550,3575,3600,3625,3650,3675,3700,3725,3750,3775,3800,3825,3850,3875,3900,3925,3950,3975,4000,8000) | params_247 |
| LUXthresholds | c() | params_247 |
| LUX_cal_constant | c() | params_247 |
| LUX_cal_exponent | c() | params_247 |
| LUX_day_segments | c() | params_247 |
| L5M5window | c(0,24) | params_247 |
| cosinor | FALSE | params_247 |
| part6CR | FALSE | params_247 |
| part6HCA | FALSE | params_247 |
| part6Window | c(start,end) | params_247 |
| includedaycrit | 14 | params_cleaning |
| ndayswindow | 7 | params_cleaning |
| strategy | 1 | params_cleaning |
| data_masking_strategy | 1 | params_cleaning |
| maxdur | 0 | params_cleaning |
| hrs.del.start | 0 | params_cleaning |
| hrs.del.end | 0 | params_cleaning |
| includedaycrit.part5 | 0.666666667 | params_cleaning |
| excludefirstlast.part5 | FALSE | params_cleaning |
| TimeSegments2ZeroFile | c() | params_cleaning |
| do.imp | TRUE | params_cleaning |
| data_cleaning_file | c() | params_cleaning |
| minimum_MM_length.part5 | 23 | params_cleaning |
| excludefirstlast | FALSE | params_cleaning |
| includenightcrit | 14 | params_cleaning |
| excludefirst.part4 | FALSE | params_cleaning |
| excludelast.part4 | FALSE | params_cleaning |
| max_calendar_days | 0 | params_cleaning |
| nonWearEdgeCorrection | FALSE | params_cleaning |
| nonwear_approach | 2023 | params_cleaning |
| segmentWEARcrit.part5 | 0.5 | params_cleaning |
| segmentDAYSPTcrit.part5 | c(0.9,0) | params_cleaning |
| study_dates_file | c() | params_cleaning |
| study_dates_dateformat | %d-%m-%Y | params_cleaning |
| overwrite | TRUE | params_general |
| acc.metric | ENMO | params_general |
| maxNcores | c() | params_general |
| print.filename | TRUE | params_general |
| do.parallel | TRUE | params_general |
| windowsizes | c(5,900,3600) | params_general |
| desiredtz | Europe/Paris | params_general |
| configtz | c() | params_general |
| idloc | 2 | params_general |
| dayborder | 0 | params_general |
| part5_agg2_60seconds | FALSE | params_general |
| sensor.location | Lower back | params_general |
| expand_tail_max_hours | c() | params_general |
| recordingEndSleepHour | c() | params_general |
| dataFormat | raw | params_general |
| maxRecordingInterval | c() | params_general |
| extEpochData_timeformat | %d-%m-%Y %H:%M:%S | params_general |
| do.anglex | TRUE | params_metrics |
| do.angley | TRUE | params_metrics |
| do.anglez | TRUE | params_metrics |
| do.zcx | FALSE | params_metrics |
| do.zcy | FALSE | params_metrics |
| do.zcz | FALSE | params_metrics |
| do.enmo | TRUE | params_metrics |
| do.lfenmo | FALSE | params_metrics |
| do.en | FALSE | params_metrics |
| do.mad | FALSE | params_metrics |
| do.enmoa | FALSE | params_metrics |
| do.roll_med_acc_x | FALSE | params_metrics |
| do.roll_med_acc_y | FALSE | params_metrics |
| do.roll_med_acc_z | FALSE | params_metrics |
| do.dev_roll_med_acc_x | FALSE | params_metrics |
| do.dev_roll_med_acc_y | FALSE | params_metrics |
| do.dev_roll_med_acc_z | FALSE | params_metrics |
| do.bfen | FALSE | params_metrics |
| do.hfen | FALSE | params_metrics |
| do.hfenplus | FALSE | params_metrics |
| do.lfen | FALSE | params_metrics |
| do.lfx | FALSE | params_metrics |
| do.lfy | FALSE | params_metrics |
| do.lfz | FALSE | params_metrics |
| do.hfx | FALSE | params_metrics |
| do.hfy | FALSE | params_metrics |
| do.hfz | FALSE | params_metrics |
| do.bfx | FALSE | params_metrics |
| do.bfy | FALSE | params_metrics |
| do.bfz | FALSE | params_metrics |
| do.brondcounts | FALSE | params_metrics |
| do.neishabouricounts | FALSE | params_metrics |
| hb | 15 | params_metrics |
| lb | 0.2 | params_metrics |
| n | 4 | params_metrics |
| zc.lb | 0.25 | params_metrics |
| zc.hb | 3 | params_metrics |
| zc.sb | 0.01 | params_metrics |
| zc.order | 2 | params_metrics |
| zc.scale | 1 | params_metrics |
| actilife_LFE | FALSE | params_metrics |
| epochvalues2csv | FALSE | params_output |
| save_ms5rawlevels | TRUE | params_output |
| save_ms5raw_format | csv | params_output |
| save_ms5raw_without_invalid | TRUE | params_output |
| storefolderstructure | FALSE | params_output |
| timewindow | MM | params_output |
| viewingwindow | 1 | params_output |
| dofirstpage | TRUE | params_output |
| visualreport | TRUE | params_output |
| week_weekend_aggregate.part5 | FALSE | params_output |
| do.part3.pdf | TRUE | params_output |
| outliers.only | FALSE | params_output |
| criterror | 4 | params_output |
| do.visual | TRUE | params_output |
| do.sibreport | FALSE | params_output |
| do.part2.pdf | TRUE | params_output |
| sep_reports | , | params_output |
| sep_config | , | params_output |
| dec_reports | . | params_output |
| dec_config | . | params_output |
| visualreport_without_invalid | TRUE | params_output |
| mvpathreshold | c() | params_phyact |
| boutcriter | 0.8 | params_phyact |
| mvpadur | c(1,5,10) | params_phyact |
| boutcriter.in | c() | params_phyact |
| boutcriter.lig | c() | params_phyact |
| boutcriter.mvpa | c() | params_phyact |
| threshold.lig | c() | params_phyact |
| threshold.mod | c() | params_phyact |
| threshold.vig | c() | params_phyact |
| boutdur.mvpa | c() | params_phyact |
| boutdur.in | c() | params_phyact |
| boutdur.lig | c() | params_phyact |
| frag.metrics | c() | params_phyact |
| part6_threshold_combi | c() | params_phyact |
| chunksize | 1 | params_rawdata |
| spherecrit | 0.3 | params_rawdata |
| minloadcrit | 168 | params_rawdata |
| printsummary | TRUE | params_rawdata |
| do.cal | TRUE | params_rawdata |
| backup.cal.coef | retrieve | params_rawdata |
| dynrange | c() | params_rawdata |
| minimumFileSizeMB | 2 | params_rawdata |
| rmc.dec | . | params_rawdata |
| rmc.firstrow.acc | 1 | params_rawdata |
| rmc.firstrow.header | c() | params_rawdata |
| rmc.header.length | c() | params_rawdata |
| rmc.col.acc | c(2,3,4) | params_rawdata |
| rmc.col.temp | c() | params_rawdata |
| rmc.col.time | 1 | params_rawdata |
| rmc.unit.acc | g | params_rawdata |
| rmc.unit.temp | C | params_rawdata |
| rmc.unit.time | POSIX | params_rawdata |
| rmc.format.time | %s | params_rawdata |
| rmc.bitrate | c() | params_rawdata |
| rmc.dynamic_range | c() | params_rawdata |
| rmc.unsignedbit | TRUE | params_rawdata |
| rmc.origin | 01/01/1970 | params_rawdata |
| rmc.desiredtz | c() | params_rawdata |
| rmc.configtz | c() | params_rawdata |
| rmc.sf | 100 | params_rawdata |
| rmc.headername.sf | c() | params_rawdata |
| rmc.headername.sn | c() | params_rawdata |
| rmc.headername.recordingid | c() | params_rawdata |
| rmc.header.structure | c() | params_rawdata |
| rmc.check4timegaps | FALSE | params_rawdata |
| rmc.noise | 3 | params_rawdata |
| nonwear_range_threshold | 50 | params_rawdata |
| rmc.col.wear | c() | params_rawdata |
| rmc.doresample | FALSE | params_rawdata |
| interpolationType | 1 | params_rawdata |
| imputeTimegaps | FALSE | params_rawdata |
| frequency_tol | 0.1 | params_rawdata |
| rmc.scalefactor.acc | 1 | params_rawdata |

Appendix B. GGIR variables used in this study.

| **Name in GGIR output** | **MX metrics** |
| --- | --- |
| AD_p99.93056_ENMO_mg_0-24hr | M1 |
| AD_p99.86111_ENMO_mg_0-24hr | M2 |
| AD_p99.65278_ENMO_mg_0-24hr | M5 |
| AD_p99.30556_ENMO_mg_0-24hr | M10 |
| AD_p98.95833_ENMO_mg_0-24hr | M15 |
| AD_p97.91667_ENMO_mg_0-24hr | M30 |
| AD_p95.83333_ENMO_mg_0-24hr | M60 |
| AD_p93.75_ENMO_mg_0-24hr | M90 |

**Appendix C.** Between group comparison of LLFDI domains.

| **LLFDI** | | **Recovery groups** | | | | ***p*-value** |
| --- | --- | --- | --- | --- | --- | --- |
| **Components** | **Domains** | **Acute (pre-fracture)** | **Post-acute** | **Extended**  **recovery** | **Long-term**  **recovery** |  |
| **Disability** | **Social Role** | 42 (36-48)* | 33 (21-38)*^€^ | 36 (32-45)*^€^ | 39 (30-46)^€^ | ***^€^ *p* < 0.01** |
|  | **Personal Role** | 59 (52-71)* | 45 (34-56)*^€^ | 54 (46-63)*^€^ | 54 (45-67)^€^ | ***^€^ *p* < 0.01** |
|  | **Instrumental Role** | 71 (61-100)* | 51 (38-65)*^€^ | 61 (49-76)*^€^ | 63 (53-79)^€^ | ***^€^ *p* < 0.01** |
|  | **Management Role** | 90 (78-100)* | 78 (67-90)*^€^ | 90 (74-100)^€^ | 90 (74-100) | ***^€^ *p* < 0.01** |
| **Function** | **Upper Extremity** | 82 (72-100)* | 74 (66-88)* | 82 (71-88) | 82 (64-100) | ***  *p* = 0.039** |
|  | **Basic Lower**  **Extremity** | 74 (60-88)* | 47 (39-61)*^€^ | 61 (50-74)*^€^ | 68 (56-86) | ***^€^ *p* < 0.01** |
|  | **Advanced**  **Lower Extremity** | 52 (32-66)* | 6 (0-28)*^€^ | 30 (11-49)*^€¥^ | 50 (25-61)^¥^ | ***^€¥^  *p* < 0.01** |

* Acute group1 versus post-acute, Extended recovery groups,

€ Post-acute group 2 versus Extended recovery and Long-term groups,

¥ Group Extended recovery group versus Long-term recovery group.

**Appendix D.** Between group comparison of MX metrics (Median (P25-P75)).

| **MX metrics** | **Recovery groups** | | | | ***p*-value** |
| --- | --- | --- | --- | --- | --- |
|  | **Acute** | **Post-acute** | **Extended**  **recovery** | **Long-term**  **recovery** |  |
| **M1** | 51.1 (34.2-70.2)* | 69.0 (50.4-86.2)*^€^ | 75.4 (59.8-111.5)*^€^ | 90.2 (66.2-120.9)*^€^ | ***** *p* <0.01,^€^ *p* <0.05** |
| **M2** | 48.6 (29.4-68.8)* | 66.8 (47.9-80.6)*^€^ | 70.9 (55.9-107.5)* | 85.3 (62.0-113.8)*^€^ | ***** *p* <0.01, ^€^ *p* =0.008** |
| **M5** | 45.8 (26.0-65.1)* | 63.1 (42.6-77.3)*^€^ | 66.1 (48.5-95.3)* | 77.7 (54.6-105.9)*^€^ | ***** *p* <0.01,^€^ *p* = 0.028** |
| **M10** | 41.2 (22.3-61.4)* | 60.4 (39.1-74.5)* | 61.8 (43.8-85.3)* | 72.7 (50.5-95.8)* | *** *p* <0.01** |
| **M15** | 38.3 (20.7-60.6)* | 60.1 (34.3-74.1)* | 60.5 (40.3-80.5)* | 69.4 (48.5-88.5)* | *** *p* < 0.01** |
| **M30** | 37.9 (17.3-59.5)* | 56.8 (29.0-71.4)* | 55.9 (36.5-73.5)* | 58.6 (38.7-79.0)* | ***** *p* < 0.015** |
| **M60** | 34.9 (14.2-59.0)* | 49.9 (25.7-69.7)* | 44.5 (28.7-64.3)* | 48.2 (27.0-67.8)* | *** *p* <0.05** |
| **M90** | 32.4 (12.3-56.9)* | 47.7 (22.6-67.7)* | 38.1 (22.7-57.6) | 43.4 (22.9-64.0) | *** *p* = 0.033** |

* Acute group1 versus post-acute, Extended recovery groups,

€ Post-acute group 2 versus Extended recovery and Long-term groups,

¥ Group Extended recovery group versus Long-term recovery group.
